# The dynamics of Covid-19: weather, demographics and infection timeline

**DOI:** 10.1101/2020.04.21.20074450

**Authors:** Renato H. L. Pedrosa

## Abstract

We study the effects of temperature, absolute humidity, population density and when country/U.S. state reached 100 cases on early pace of Covid-19 expansion, for all 50 U.S. states and 110 countries with enough data. For U.S. states, weather variables show opposite effects when compared to the case of countries: higher temperature or absolute humidity imply faster early outbreak. The higher the population density or the earlier the date when state reached 100^th^ case, the faster the pace of outbreak. When all variables are considered, only population density and the timeline variable show statistical significance. Discounting the effect of the timeline variable, we obtain an estimate for the initial growth rate of Covid-19, which can be also used to estimate the basic reproduction number for a region, in terms of population density. This has policy implications regarding how to control the pace of Covid-10 outbreak in a particular area, and we discuss some of them. In the case of countries, for which we did not have demographic information, weather variables lose statistical significance once the timeline variable is added. Relaxing CI requirements, absolute humidity contributes mildly to the reduction of growth rate of cases for the countries studied. Our results suggest that population density should be employed as a control variable and that analysis should have a local character, for subregions and countries separately, in studies involving the dynamics of Covid-19 and similar infectious diseases.

## Introduction

The first reported cases of what was later denominated Covid-19, caused by new virus of the group *Coronaviridae*, occurred in China during the last month of 2019. Cases soared during the second half of January, going from 80 on January 18^th^ to over 9,000 by the end of the month, when there were 14 cases reported in Japan and only a few other across Asia and only 7 in the United States. By mid-February, there were cases in many other countries, but still in relatively small numbers, and none in Africa or South America, which have most of their territories in the Southern Hemisphere, then under Summer. By the end of that month, some countries, besides China, were already facing an epidemic situation. The first positive case reported by a South American country occurred on Feb. 26^th^, in Brazil. Since then, the virus started to spread, slowly, with all cases being of people arriving from other countries, especially from Italy, or who had direct contact with them. By March 6^th^, it was recognized that local untraceable transmission was occurring in Brazil and the number of cases grew rapidly. As we will see, the pattern, for the 10 days starting on the day of the 100^th^ case, in Brazil as in many other countries, is exponential to a high degree, with varying growth rate values. During the whole period since the start of the pandemic until the end of March, weather in Brazil has been warm and humid, with temperatures frequently soaring above 32C and absolute humidity never below 10g/m^3^. This raised the possibility that warm and humid weather will not help contain the spread of the virus, as is typical of many viral diseases (*1,2,3,4,5*). There are various studies with early, somewhat conflicting, results on those relationships for Covid-19 (*6,7,8,9,10,11,12,13*), as discussed in the NASEM report (*14*). This study proposes to expand the analysis to include, besides weather information, demographic (population density) and the timeline of outbreak (date of 100^th^ case) as variables. The idea that higher population density would increase the growth rate of Covid-19 is expected (*15*), the research we are reporting helps understand how e by how much.

Besides weather, there has been also a wide range of estimates of the basic reproduction number *R_0_* of Covid-19, the average number of people infected by an infectious person, from 2 to over 6, not including CI’s, all based on the early data from China (*16*). We will see that population density must be taken into account and may help explain the variability of *R_0_*, as it impacts the estimate of early daily growth rate of the disease, used in many of those estimates.

We develop multivariable linear models in which the dependent variable is the early daily growth rate of Covid-19, which has been estimated by the coefficient of the best exponential fit to the evolution curve of cases in the period of 10 days starting on the date when region reached the 100^th^ case. It will be denoted by *k*. The control variables are: average temperature and absolute humidity values during the 25 days starting 15 days before the region reached the 100^th^ case; the date when the 100^th^ case occurred; and the (log of) population density for the counties with higher participation in the spread of Covid-19 (for U.S. states only). We find that weather variables, albeit significant in some of the single-variable models (but with opposite effects for the two groups of regions, U.S. states and countries), lose significance when the timeline and/or demographic variables are introduced. In the first case, for states and countries, the later the date of the 100^th^ case, the slower the initial pace of expansion of Covid-19. In the second one, the higher the population density U.S. states, the faster the disease has spread in its initial phase. The model with both variables, only available in the case of U.S. states, furnishes the best estimates, explaining 71% of the variability of the early daily growth rate *k*. In the case of countries, if one relaxes statistical requirements, absolute humidity (together with date when country reached the 100^th^ case) shows a small positive impact on the early daily growth rate of Covid-19, but we do not expect even that small effect to remain once one includes population density and possibly other variables in the model.

Discounting the effect of the date of 100^th^ case, we obtain an estimate of the initial daily growth rate of cases of Covid-19 as a function of population densities of U.S. states. The relation is given by

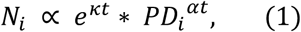

where *PD_i_* is the population density of region, *κ*, *α* are universal constants (not depending on region), which are derived from our analysis. A similar model should hold for regions in other countries, which may be verified by further studies, possibly with different parameters. It would also help explain part of some unexpected behavior among countries and among subregions and cities within a country and would be useful for policy decisions regarding how to develop adequate social distancing measures. We end our study by developing a preliminary relationship between the basic reproduction number of Covid-19 and population density, derived from results of models developed in our analysis.

The timeline variable, the data when the region reached the 100^th^ case, is likely a proxy for early social distancing practices, even before government measures are put in place, so that it actually depends on how the population perceives the dangers of the infection and how governments respond to it, sometimes even before the first cases had been detected, but after the infection had shown that it could become a health issue. It may also be related to other societal variables, such as number of persons per dwelling (crowding), intensity of use of mass public transportation, income, education, etc. Population density is an independent variable that impacts the general rate of expansion of Covid-19 and, possibly, if lower, makes distancing measures easier to implement and maintain, helping the control of its growth rate.

## Data and methods

### Confirmed Covid-19 cases

Databases of reported cases of Johns Hopkins University’s Center for Systems Sciences and Engineering (CSSE/JH) (*17*) for U.S. states, of the European Centre for Disease Prevention and Control (ECDC/EU) (*18*) for countries.

### Weather data and averages of temperature and absolute humidity

NOAA Integrated Database (ISD) (*19*) of meteorological observations, through the R package “worldmet” (*20*). For countries, we used the station nearest to the capital with 100% coverage, when available. In the case of the United States, data for the main airports in Seattle, San Francisco and New York City. For U.S. states, the data from main airport in the largest city in the state. For Brazil, data for the main airports of São Paulo and Rio de Janeiro. From NOAA weather data, the absolute humidity *AH* (g/m^3^) is computed from temperature *T* (C) and relative humidity *RH* (%) using an approximation formula derived from the Clausius-Clapeyron equation (*8,21,22*),

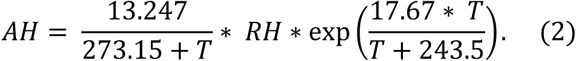

The averages for both variables were computed during the period of 25 days starting 15 days before the day the region reached the 100^th^ case, covering a span of two weeks before confirmation and the next 10 days used to estimate the exponential coefficient for the evolution of cases. The above equation shows that AH depends exponentially on T, to a high degree, for constant RH, in the usual range of values of temperature. This may be seen in Fig. 6 and will have impact on our models.

### Countries and U.S. states

We have considered the 110 countries and that had at least 10 days of data starting when they had reached the number of 100 accumulated cases, end-date being at most April 10^th^. All 50 U.S. states had already reached that stage by that date.

### Population density of U.S. states and counties

All data used is for 2019 (*23,24*). We tested three variables linked to county population density in the U.S.: density of the densest county and (weighted) average of densities of counties which responded for at least 60% of the cases in a state, in both arithmetic and geometric versions. The last two cases provided the best fitting, very close in results. Regarding the density of densest county in a state, there are too many exceptions where they are not the main contributor to the early spread of Covid-19. For example, in California, Santa Clara, San Mateo, San Diego and Los Angeles had higher early numbers of cases compared to San Francisco, the densest county in the state. Similarly, Westchester county was a very important contributor to the number of cases in New York State, later surpassed by New York City county, but it is over 30 times less dense than the latter (867 to 27,775 pop/km2, respectively). We will see how that is related to the growth rate of Covid-19 in those two areas. After some testing, we decided to use the arithmetic weighted averaging of densities, i.e., the usual aggregate density which is computed by the ratio between total population and total land area of all counties involved, as it provided slightly better fitting properties in relevant models.

We have employed as control variable the natural logarithm of density. Density enters econometric analysis via its log-transformation (*25,26*) and it is also typical to apply log to variables of concentration in chemistry, like in the computation of pH, and in other fields. In health-related studies, it is also usual to use logarithm of density (*27*). In epidemiology, carefully modelling the dependence of the basic reproductive rate (*R_0_*) of infectious diseases on population density (*28*) shows that one must use non-linear scaling when dispersion is high, which is the case here, as population density is concentrated on smaller values (Fig. 1.f). As we analyzed various options of transformations, both in terms producing a more evenly spread distribution of the variable and of our model fitting, it turned out that the *log*-transformation of population density showed not only good distribution properties (Fig. 1.g), but proved to be well adapted as a control variable in the models.

**Figure 1.a-g.**
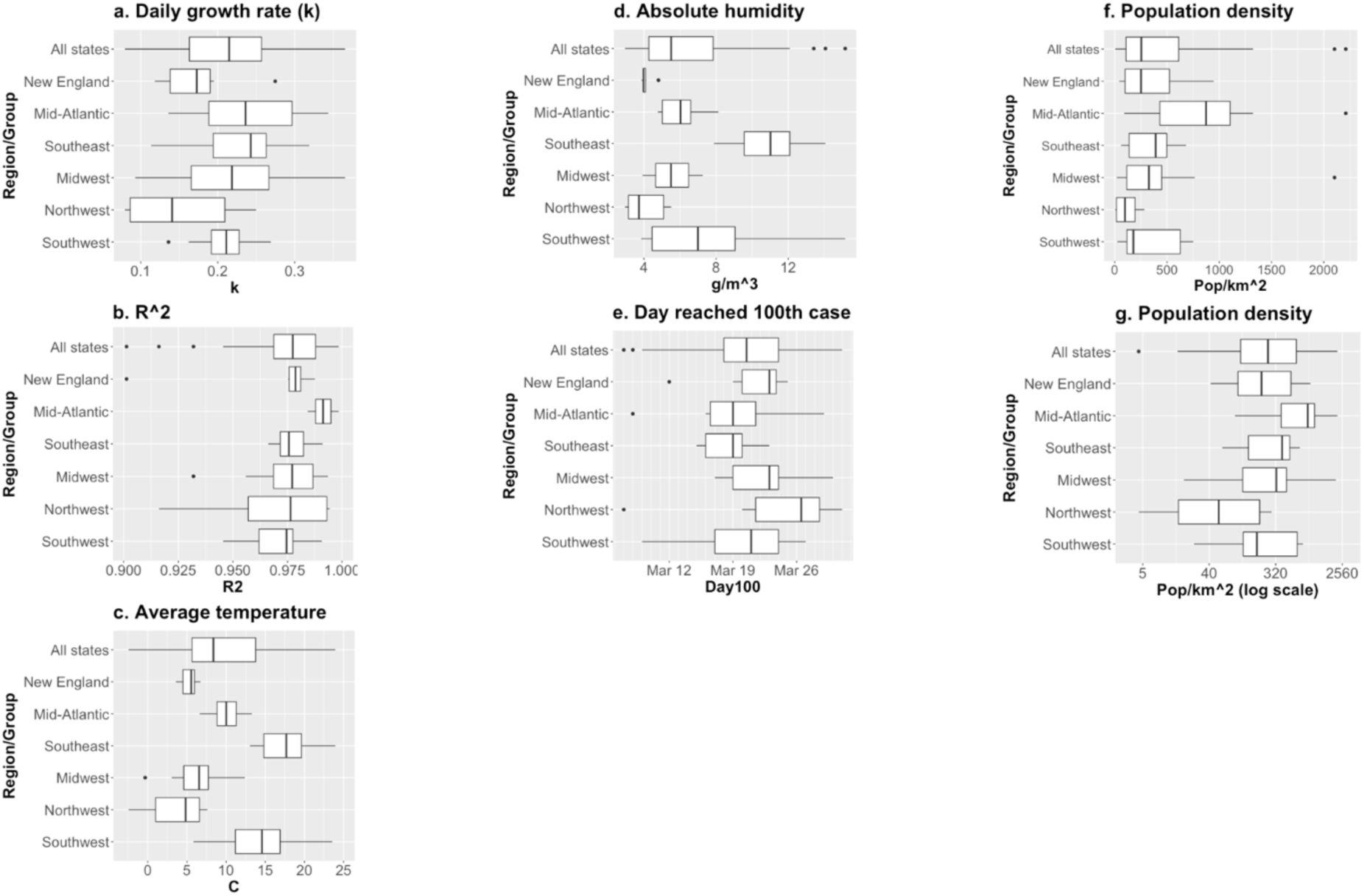
Boxplots for regression variables and for *R^2^* for the exponential regression for *k* (Eq. 3). U.S. states, by region. Data: CESS/JHU, NOAA/USDC, U.S. Census (2010).

### Growth rate of number of cases

We considered the period of 10 days starting on the day the number of cases reached 100 and computed a simple linear regression for *log(N_i_)* as function of day (*t*), where *N_i_* is the number of accumulated observed cases for region *i* at a given day of the period, given by the model

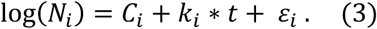

The growth rate variable, the dependent variable in the models we develop, is the estimated *k_i_*, the initial daily growth rate of cases of Covid-19. We have also checked 12-day and 15-day windows starting on the 100th-case day, but there was no relevant impact on the results of the models. We preferred the 10-day window as it provided the best fitting parameters for models given by Eq. 3. We have also estimated the growth rate using endpoints, but we wanted to be able to check the fitting of evolution curves to an exponential and if that affected results. Also, sudden jumps in the reported number of cases cause similar jumps of endpoints estimate, while the regressed estimate follows a smoother path. The choice of day of 100 accumulated cases is related to when it is expected that local transmission would be under way, as that is when an exponential increase is expected to start. When states/countries reach 100 cases, there are already a few communities with at least 20 cases; it has been estimated that when a U.S. county has reaches 20 cases, the chances that there is ongoing local transmission are 99% (*29,30*). For example, for the United States as a country, the date of the 100^th^ case was March 3^rd^, when Washington and California led the country, with 27 and 25 cases, respectively, concentrated in King County and counties around San Francisco, respectively. Within the 10-day period used in the estimation, New York had also become a hotspot of cases, and on the last day of the 10-day period, it was second in number of cases, and other states had surpassed 20 cases.

### Hypothesis on k_i_

For the estimate *k_i_* to be representative of the speed of transmission of the novel coronavirus in a community we need, first, that *the way the cases were accounted for did not change significatively along the 10-day period used for the models*. The evidence from the estimates corroborates that assumption, as the values of *R^2^* for regressions (1) are mostly above 0.95 (Figs. 1.b, 2.b); secondly, that *the number of positively tested cases of people with various levels of symptoms, or of hospitalized cases, are constant fractions of the total number of people infected (of which, many, are asymptomatic)*. Some countries have tested all people with any symptom, others only those hospitalized (the case for Brazil), and others with in-between levels of symptoms. The estimated *k_i_*, under those two hypotheses, would not depend on the alternatives considered by different countries (or states) regarding how they measured confirmed cases.

**Figure 2.a-e.**
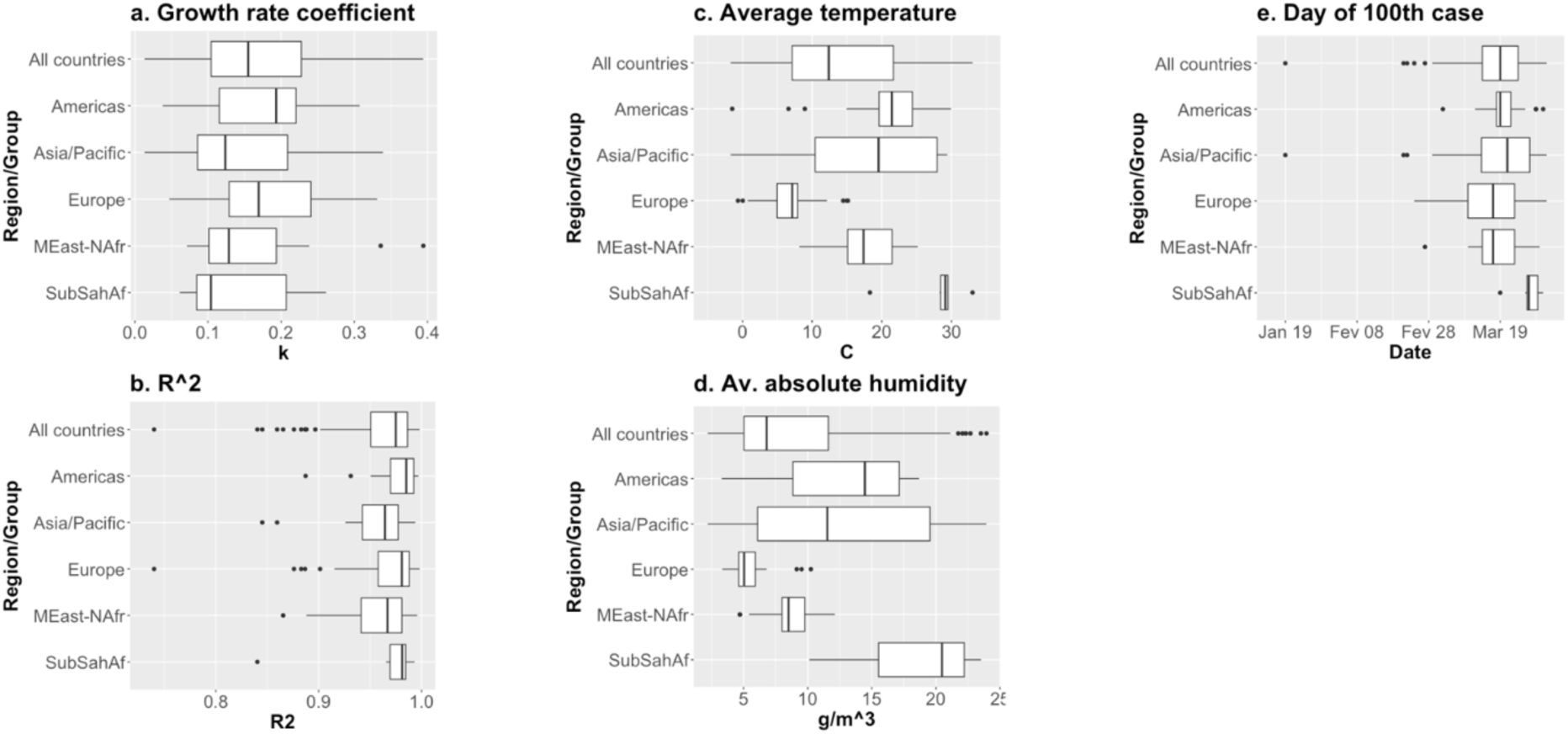
Boxplots for regression variables and for *R^2^* for the exponential regression for *k* (Eq. 3). Countries, by region. Data: ECDC/EU, NOAA/USDC.

### Modelling the early dynamics of Covid-19

Using the data described above, we consider the following linear regression model for the U.S. states:

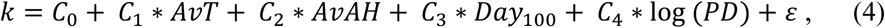

where *k* is the exponential growth rate coefficient estimate (Eq. 3), *AvT* is average temperature, *AvAH* is average absolute humidity, *Day_100_* is the date when state reached the 100^th^ case and *PD* is the population density variable described above. For the collection of 110 countries we use the same model without the last regressor. For both cases, average temperature and absolute humidity show high levels of association (Fig. 6), so that we have to be careful when using both variables in the same model. We will further discuss those points in the variables and modelling sections below. Before we move on to analyzing the data and models, we observe that we have also considered another timeline variable, given by the number of days between the occurrence of the 1^st^ and the 100^th^ case, but it did not introduce any new statistically relevant insight into the analysis, so we have opted to leave it out in the final version of the paper.

*Software*: *R*, *worldmet* package.

## Preliminary analysis

### Variables

The dependent variable given by the initial daily growth rate *k* shows appropriate distribution properties, which may be seen from Figs. 1.a and 2.a. That allows us to go forward with traditional linear regression models, without need for transforming that variable or using GLM. We see that *k* is spread along a very wide set of values, from almost 0 to 0.4. For countries, mean = 0.169, median = 0.155 and SD = 0.082. For U.S. states, mean = 0.209, median = 0.215 and SD = 0.068. Fig. 3 has the evolution of *k* for the four states with earliest date of the 100th case, which have some outlier behavior regarding our models, as will be explained later, California, Massachusetts, New York and Washington. We observe that the first three states in that group show a peak for the values of *k* that do not coincide with date which we used to start the 10-day window for calculating the value of *k* for that state. This will be further discussed when we develop our models to isolate the effect of population density on the early daily growth rate. Washington shows a pattern which is similar to that of most other states, with a monotonously decreasing trend for *k* from the start, which would seem to be associated to some early attenuation of growth, even though it was the first state to reach 100 cases.

**Figure 3.a-d.**
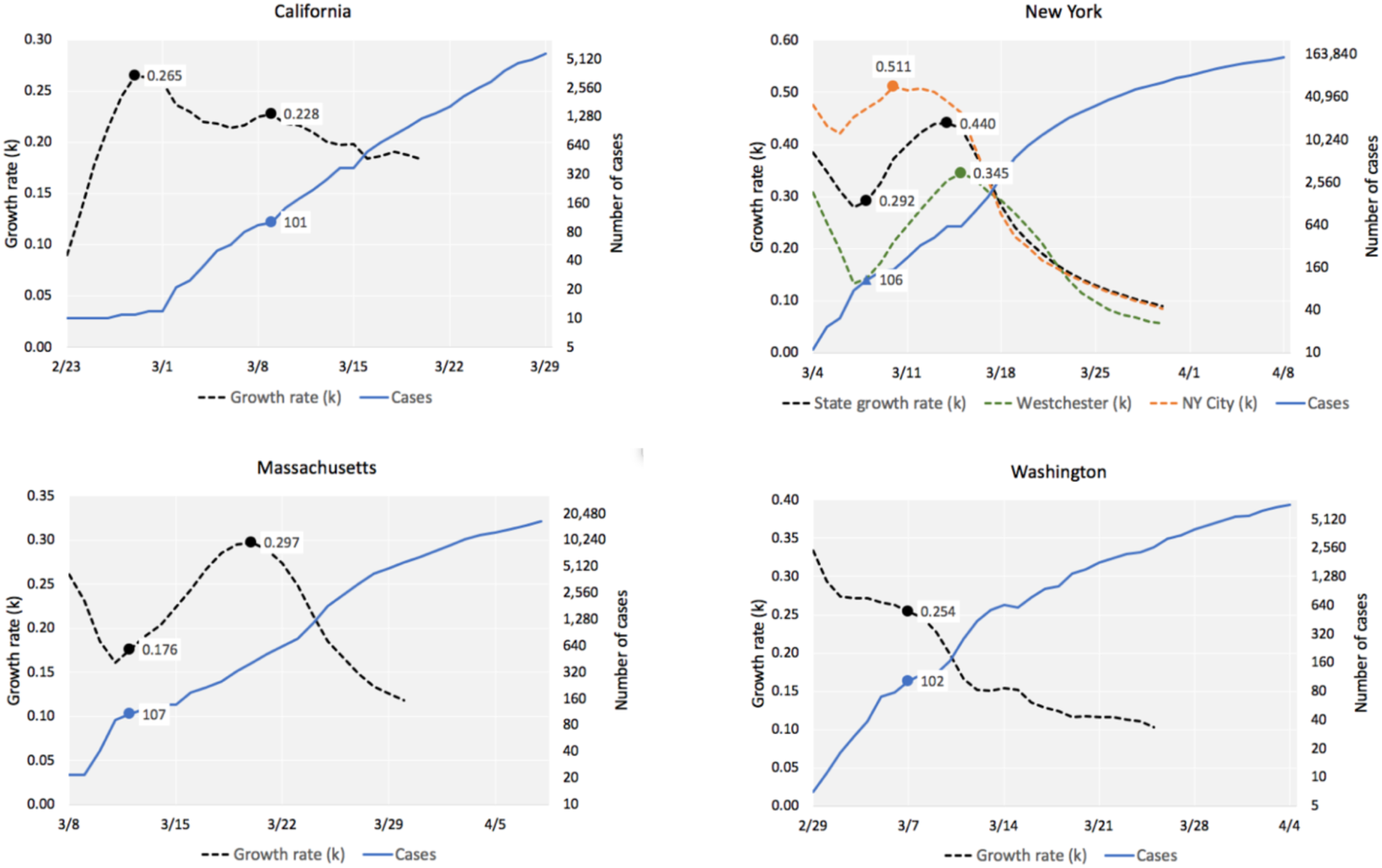
Number of cases and evolution of *k*, computed using a 10-day moving window. States with early outbreaks (100^th^ case on or before March 12^th^). Values of *k* marked on the first day of 10-day window. Graph for New York includes the two counties with most cases in the 10-window period used to compute the estimate of *k* for the state. All graphs include the points for date when state reached the 100^th^ case. Point of maximum also annotated for *k* curves. Data: CSSE/JHU.

The values of *R^2^* for regressions employed to estimate *k* (Eq. 3) were typically above 0.90 and most above 0.95 (Figs. 1.b, 2.b). In the case of U.S. states, *R^2^* > 0.90 in all cases. For countries, there were a few cases for which the regressions following Eq. 3 with *R^2^* below 0.90. To visualize how well the exponential model for the growth rate *k* fits the actual trajectories, Fig. 4 displays the exponential prediction and the curves for actual accumulated number of positive Covid-19 cases, for selected countries, including eight of the eleven countries that had *R^2^* < 0.90 and another group with *R^2^* > 0.95. It is interesting that there is little qualitative difference between the two groups of countries, except for the case of Denmark, which had *R^2^* = 0.74, the lowest among all countries. From the group of those countries with relatively low values for *R^2^*, it seems that lower values of *k* tend to come from cases with lower *R^2^* as well, but we saw no such trend for all values of *R^2^*, so we decided not to restrict the set of countries in the study based on that variable. Testing the models without Denmark did not brought up any new statistical behavior, so we kept it in all models.

**Figure 4.**
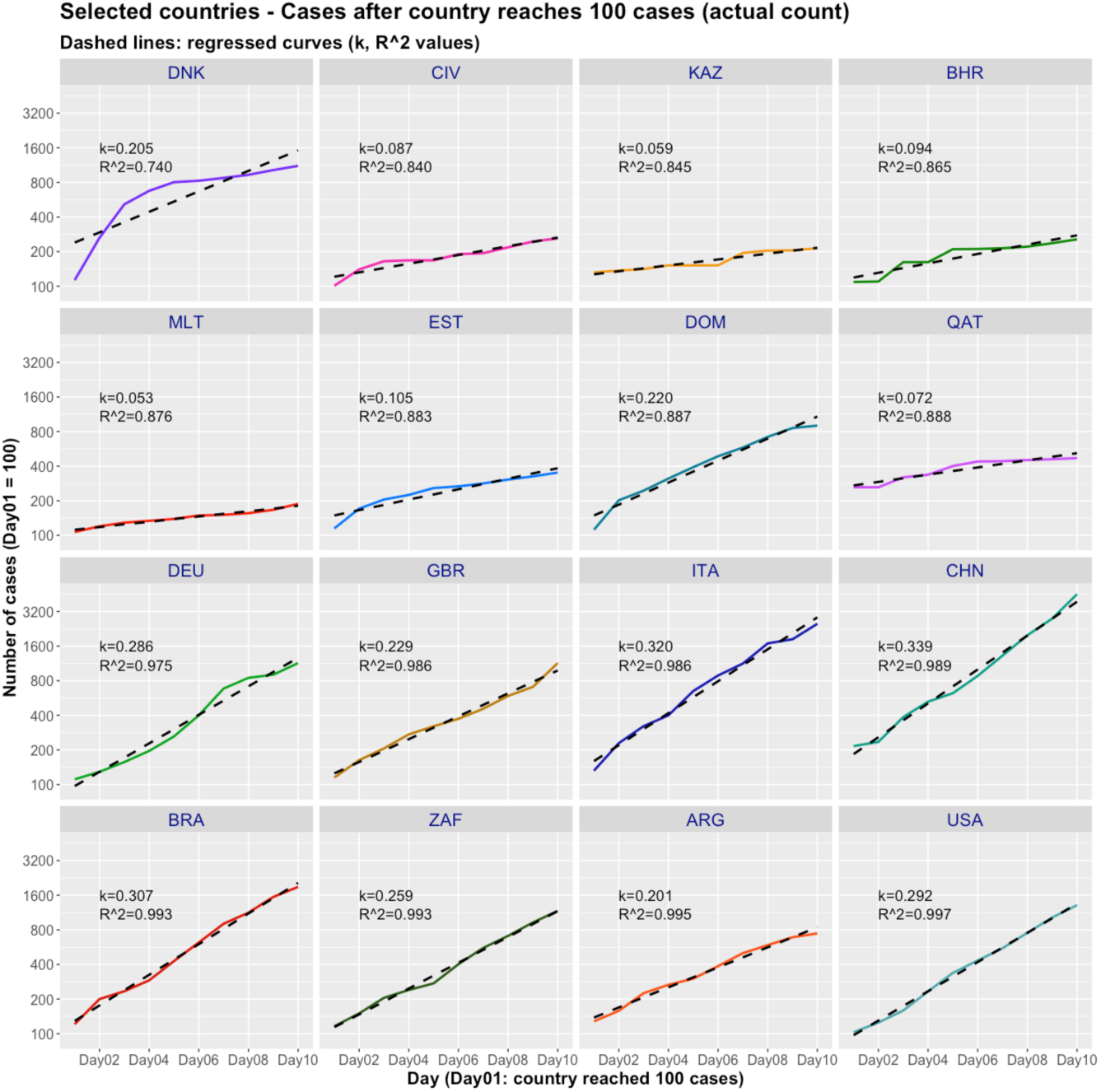
Actual and regressed (Eq. 3) curves of number of confirmed cases, selected countries, 10 days starting on the day of 100^th^ case. Values of *k* and *R^2^* for the regressed exponentials. Data: ECDC/EU.

Figs. 5.a-d show the daily behavior of average temperature and absolute humidity, along the 25-day periods used to estimate the initial growth rate for U.S. states and countries. Figs. 6.a/b show that there is strong exponential association between the two variables. The squares of correlation for the exponential trend lines are above 0.85 in both cases, and with a few outliers removed, above 0.89 for countries and above 0.92 for U.S. states (shown in the graphs). This is expected, as Eq. 2, for constant relative humidity, is closely approximated (*R^2^*>0.99) by an exponential of temperature in the usual natural range of values of temperature. For linear trend lines, *R^2^* is above 0.80 in all cases. These results imply that we should be careful when employing both variables in the same model, as we will see below, in the modelling section. Figs. 7.a-d present how temperature and absolute humidity relate to when state/country reached the 100^th^ case. We see they follow opposite trends for the two groups, a fact that will help explain I part why the timeline variable, when added to models, reduces the impact of the weather variables.

**Figure 5.a-d:**
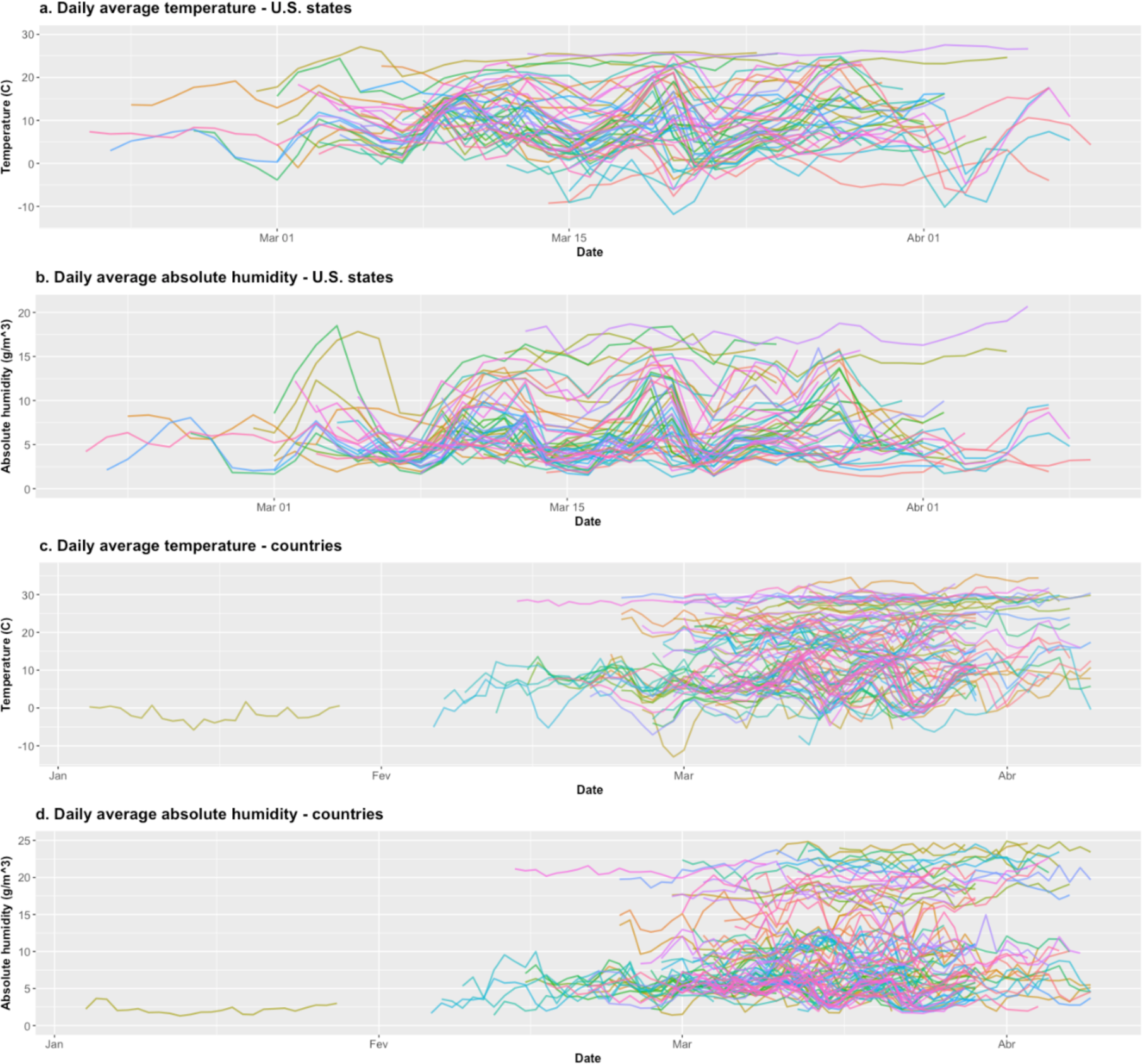
Daily average temperature and absolute humidity, U.S. states and countries. Period of 25 days, starting 15 days before state/country reached 100^th^ case. Data: NOAA/USDC, CSSEC/Johns Hopkins, ECDC/EU.

**Figure 6.a-b.**
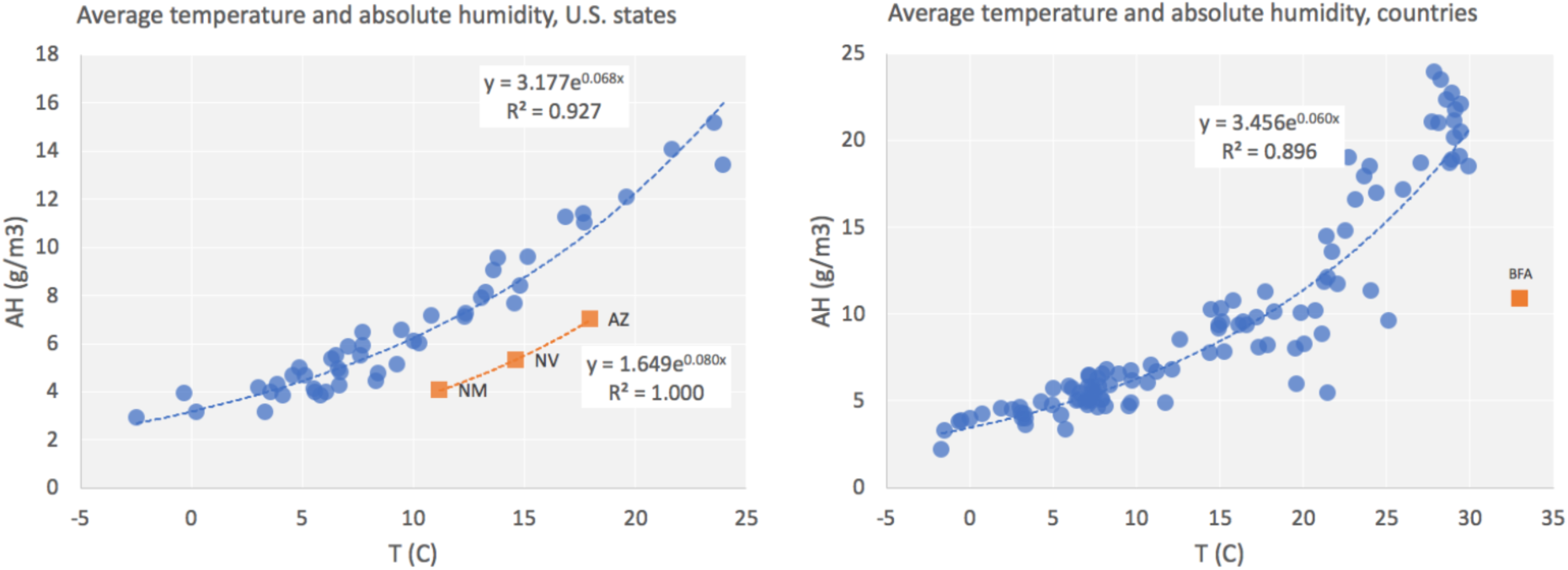
Average temperature and absolute humidity, exponential trend lines: a. U.S. states, b. countries. Parameters and trend lines have excluded states and country in orange (Burkina Faso) (see text). Data: NOAA/USDC.

**Figure 7.a-d.**
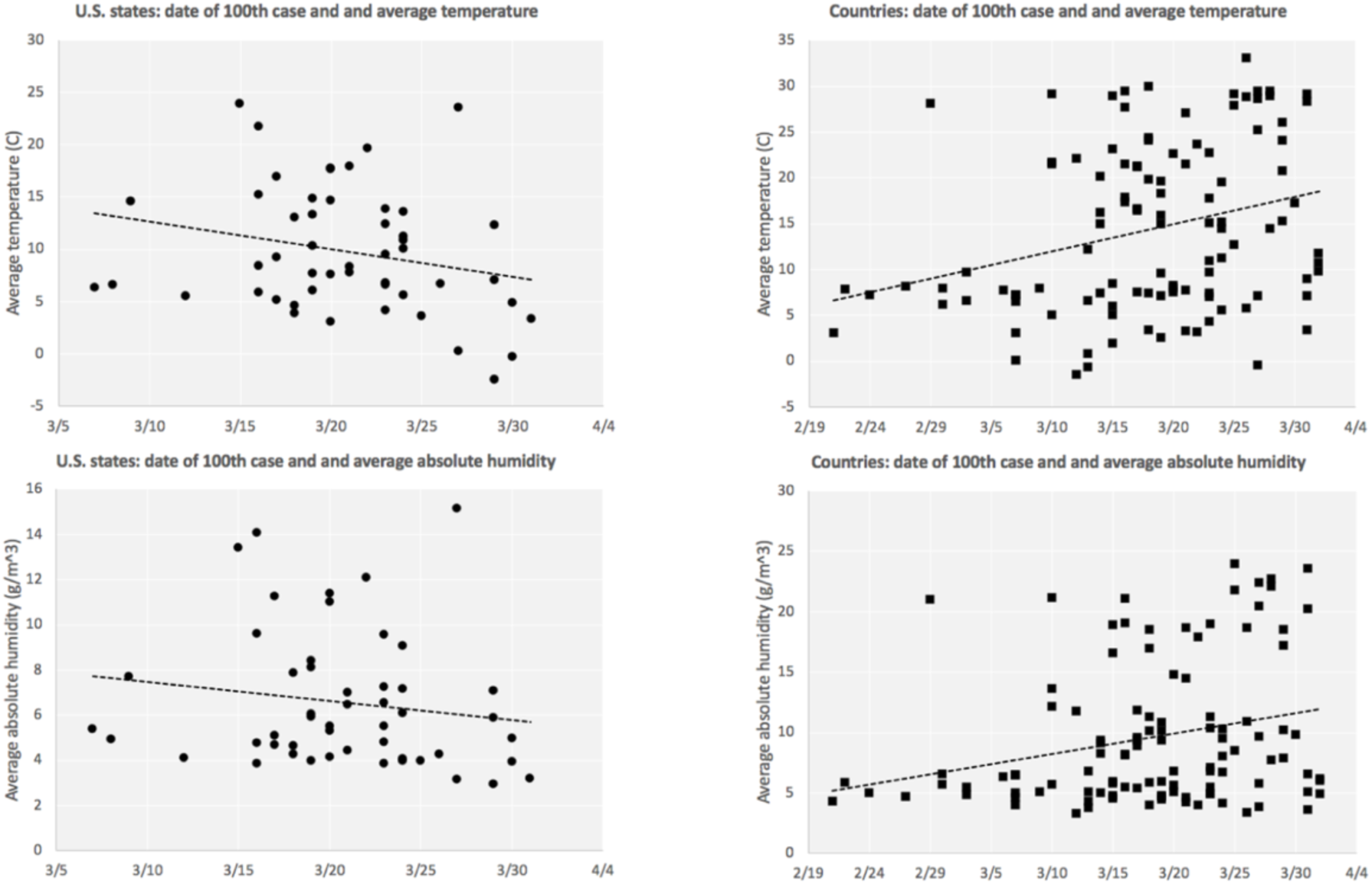
Scatter plots of weather variables and date of 100^th^ case, U.S. states and countries. Data: CSSE/JHU, ECDC/EU, NOAA/USDC.

**Figure 8.**
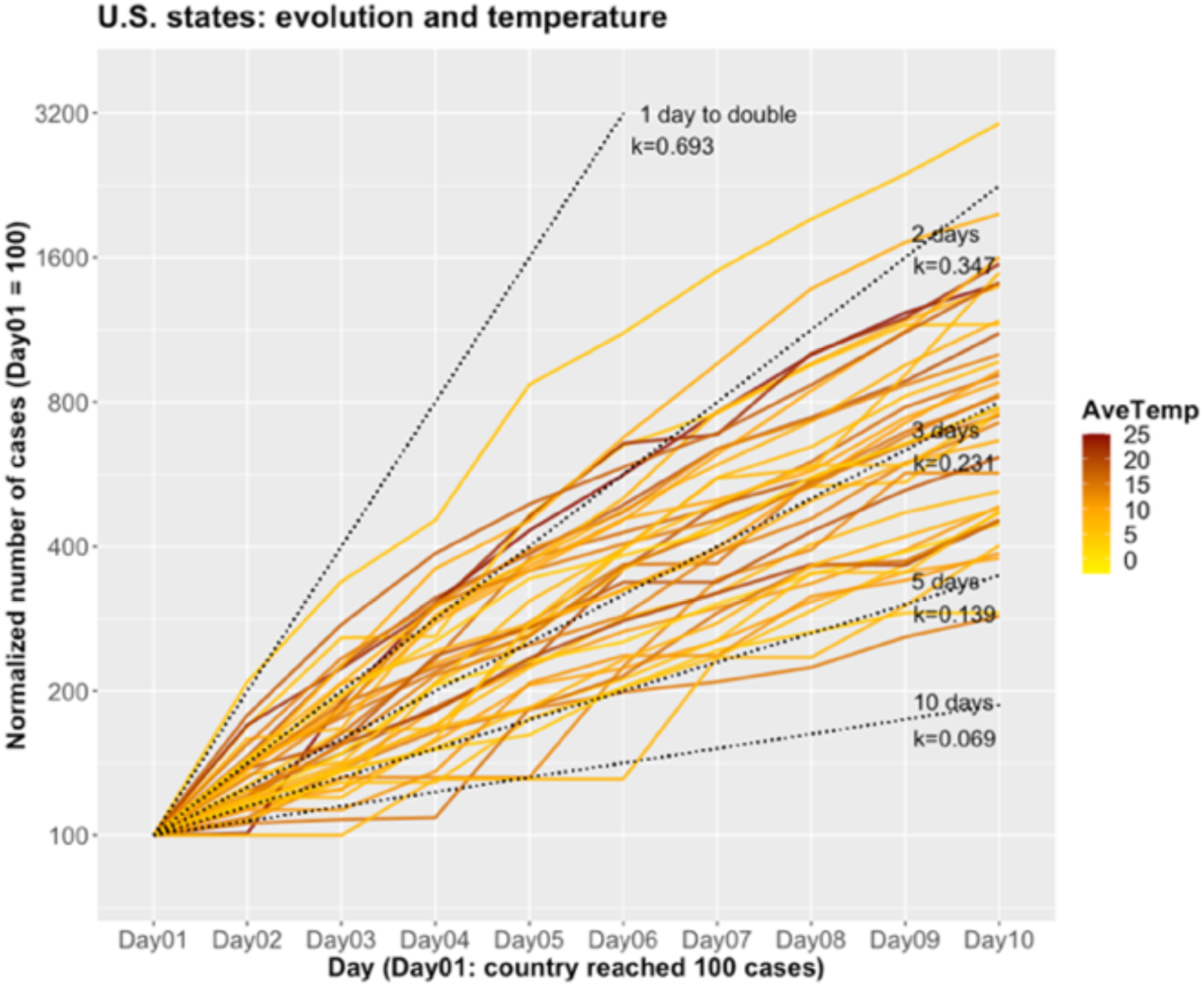
Number of confirmed cases for 10 days starting when U.S. state reached 100 cases (normalized count, Day01 = 100), by temperature. Dotted lines: exponential curves with number of days to double the number of cases and value of *k*. Data: CSSE/JHU, NOAA/USDC.

For the date when state/country reached the 100^th^ case, China, among countries, shows a strong outlier behavior, as it did so on Jan. 19^th^, more than a month before any other country (Fig. 2.e). We present models with and without China, whenever that variable was involved, even though the impact of that was minimal. In the case of the variable of date when state reached the 100^th^ case, there are four states (California, Massachusetts, New York and Washington) which showed outlier character, as they were the first to reach that stage (seen clearly in Fig. 9.c, but also in Fig. 10). Removing those four states did not change the overall behavior of models, but improved estimates in all models in which it was included. For all quantitative analyses we have employed the models with the restricted set of states, omitting those four states.

**Figure 9.a-d.**
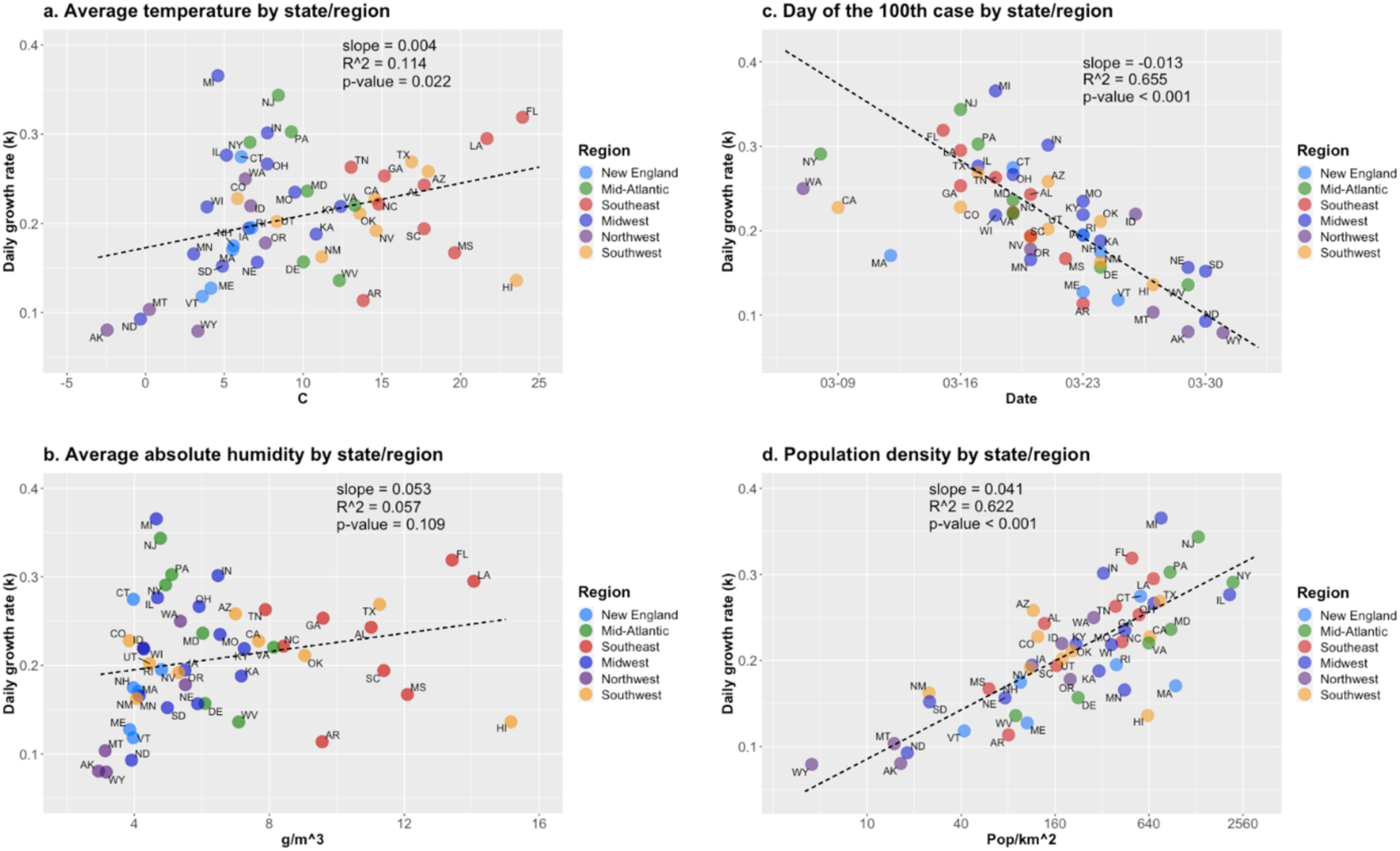
Scatter plots of exponential growth rate (*k*) and control variables, U.S. states. Regression parameters and line omit CA, MA, NY, WA (see text) Data: CSSE/JHU, NOAA/USDC, U.S. Census (2010).

**Figure 10.**
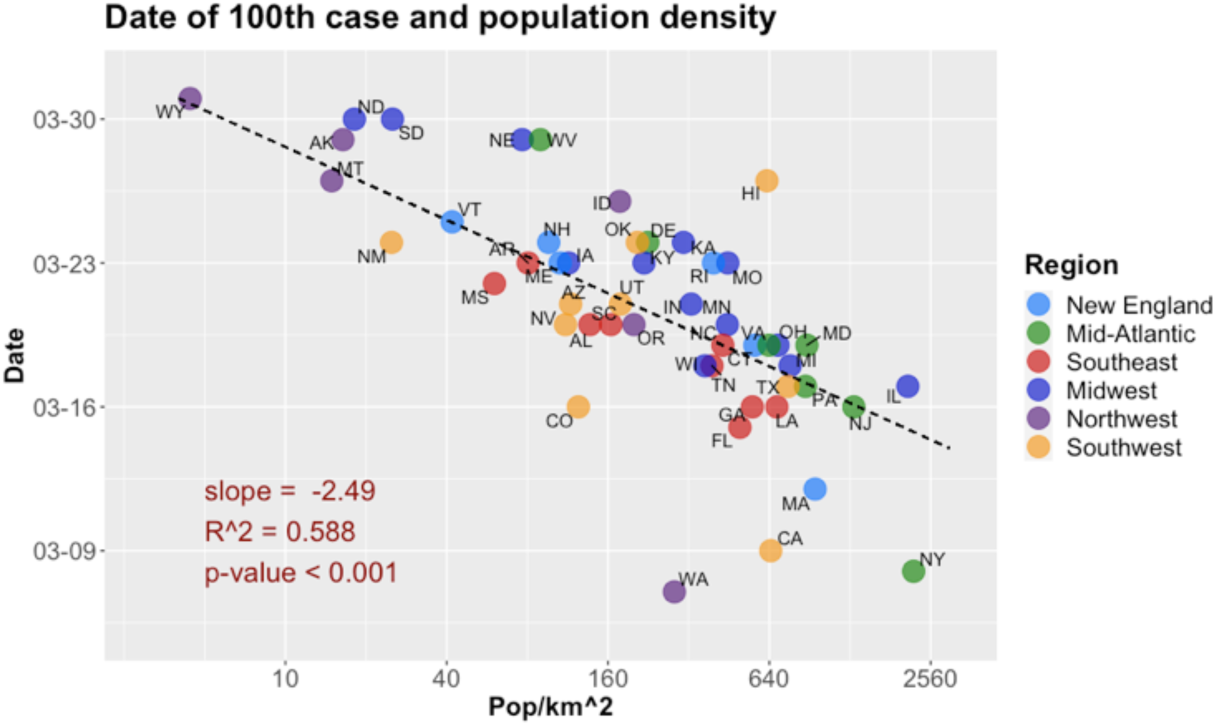
Scatter plot of date of 100^th^ case and population density, U.S. states. Regression parameters and line omit CA, MA, NY, WA (see text). Data: CSSE/JHU, U.S. Census (2010).

Population density was used in log-transformation, as already discussed. We recall that density was computed as the aggregated density of counties which contributed with at least 60% of the cases along the 10-day window used to estimate the daily growth rate *k*, for each state. There was only one exception, that of Idaho. Using the above criterion, data from two counties would be employed, Ada and Blaine. Ada is the densest county in the state, with a population of 482,000 and density of 178 pop/km^2^, while Blaine is a mountain tourist area with 23,000 population and density of only 3.4 pop/km^2^. As of Apr. 10^th^, Blaine had 19.4 cases per 1,000 pop., Ada, 1.03, Canyon, 0.60, and Kootenai, 0.25. Because of that disparity, and the special situation of Blaine County, which had in the tourist inflow a probable cause of the high intensity of infection, we decided to use density of Ada County as the density value for Idaho. Overall, that choice did not change the estimates and models, but, when we later adjust growth rate by day of 100^th^ case and then compute the level of expected daily growth rate for Idaho in terms of density, that would impact the state’s final estimate in a significant way.

Fig. 8 shows the evolution curves of cases for U.S. states for the 10-day period used to estimate the growth rate *k*, by temperature. It shows that there is a wide range of values of *k*, and no obvious trend in the temperature averages w.r.t. *k*. The analogous graphs for absolute humidity, and for countries, not included, are similar.

## Models: technical aspects

Models based on Eq. 4 are presented in Tables 1 and 2 for U.S. states and countries, respectively. The intervals for coefficients use 95% CI. We omit the values of intercepts, as they are not relevant for this part of analysis. We include *F*-statistic *p*-values for the multivariable cases and Shapiro-Wilk test results for the residuals of all relevant regressions. In the case of multivariable regressions, *R^2^* is the adjusted value.

**Table 1.**
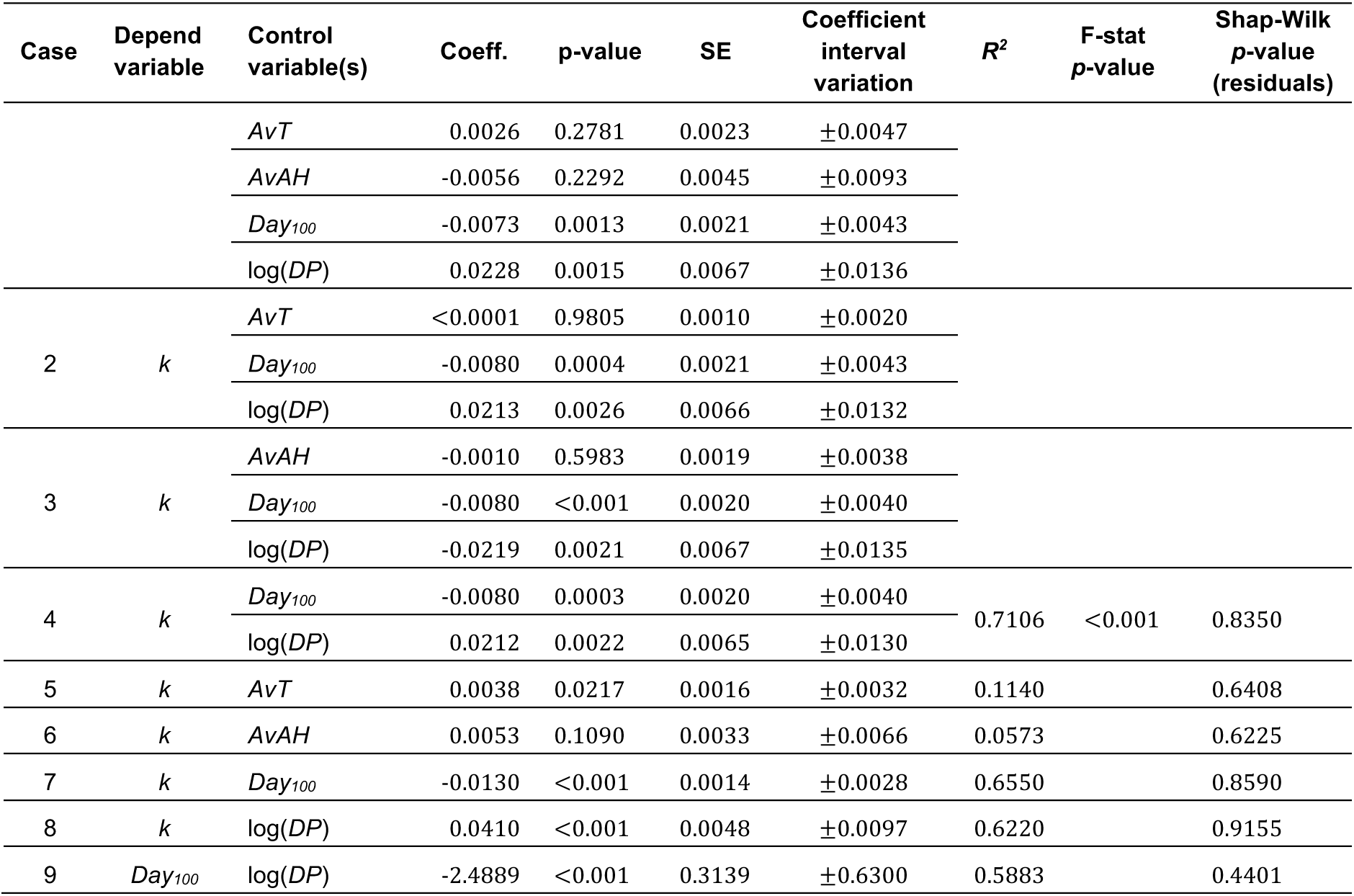
Regression models for U.S. states. All models exclude CA, MA, NY, WA (see text). Coefficient interval: 95% CI estimate. *R^2^* for model 4 is the adjusted value.

### U.S. states

Table 1 presents the results of regressions for the exponential growth constant *k* and regressions for timeline variables w.r.t. population density for U.S. states. As the qualitative behavior of models employing all states is the same as that excluding the states which have outlier behavior in relation to the regression of k w.r.t. the timeline variable of data of the 100^th^ case, and all regressions with this restricted set show sharper estimates, we have presented only those models. Model 1 shows that only the date of the 100^th^ case and density have statistical significance in the complete model. Since it includes both weather variables, and they could be interacting and increasing their p-values, given their high level of collinearity (discussed above), we have Models 2/3 with only one of them included. Again, the weather variables show high p-values. Model 4 is the only multivariable model for which all variables have coefficients rejecting the null hypothesis and will be the basis for discussion developed below.

Model 9 present the association of the relevant timeline variable and population density. Even though the association is very intense, one must note that both variables survived in Model 4, indicating that there are effects of each which cannot be explained by the other, regarding impact on the early daily growth rate of Covid-19.

### Countries

Models 1/1^†^ in Table 2 show that the variable *Day_100_* is the only one statistically significative in the complete model. Model 1^†^ omits China, as explained earlier. The 1-variable models show p-values below 0.05, but one must note that the residuals, in the cases of the weather variables, both cases, have low p-values in the Shapiro-Wilk test. An inspection of scatter plots in Figs. 11.a-b does not provide obvious choices of countries that would be generating the non-normality of the residuals. Model 2, for *Day_100_* and *AvAH* as control variables, may be considered relevant, once the CI requirement is relaxed, as we discuss below.

**Figure 11.a-c.**
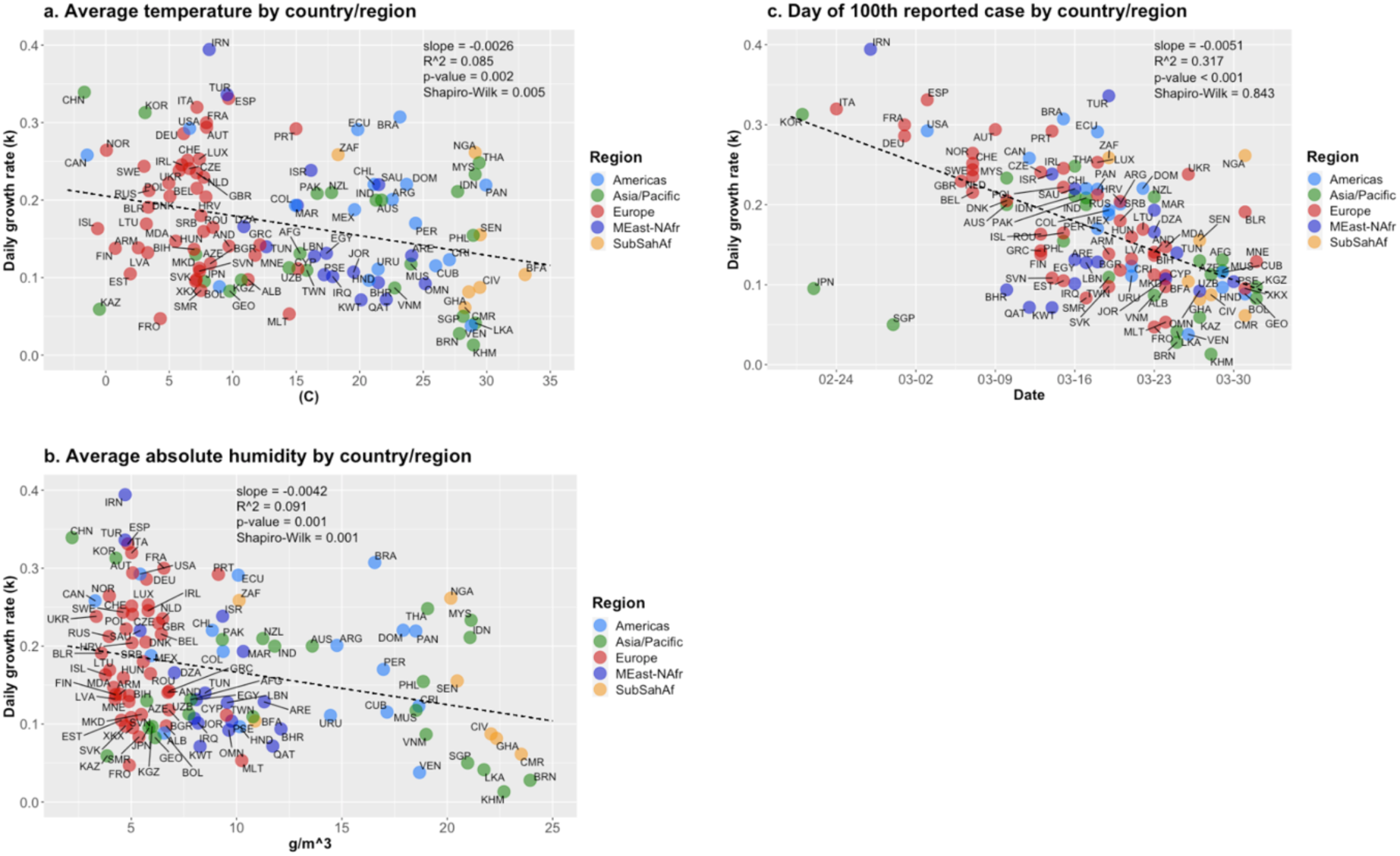
Scatter plots, exponential growth rate (*k*) and control variables. Countries. Plot c. omits China (see text). Data: ECDC/EU, NOAA/USDC.

**Table 2.**
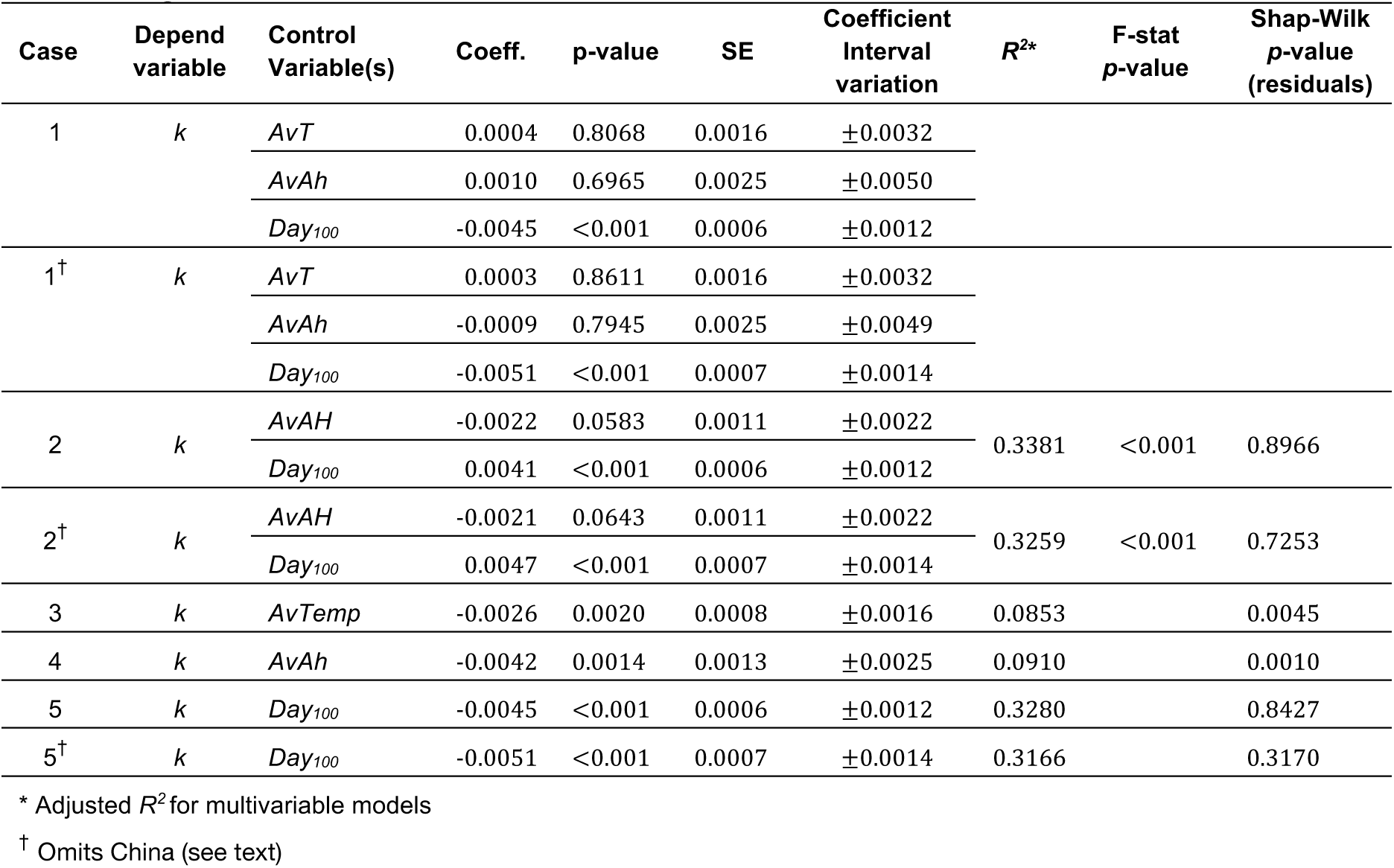
Regression models for countries. Coefficient interval: 95% CI estimate.

| Case | Depend variable | Control Variable(s) | Coeff. | p-value | SE | Coefficient Interval variation | $R^{2*}$ | F-stat p-value | Shap-Wilk p-value (residuals) |
| --- | --- | --- | --- | --- | --- | --- | --- | --- | --- |
| 1 | *k* | *AvT* | 0.0004 | 0.8068 | 0.0016 | $\pm 0.0032$ | | | |
| | | *AvAh* | 0.0010 | 0.6965 | 0.0025 | $\pm 0.0050$ | | | |
| | | *Day<sub>100</sub>* | -0.0045 | <0.001 | 0.0006 | $\pm 0.0012$ | | | |
| 1<sup>†</sup> | *k* | *AvT* | 0.0003 | 0.8611 | 0.0016 | $\pm 0.0032$ | | | |
| | | *AvAh* | -0.0009 | 0.7945 | 0.0025 | $\pm 0.0049$ | | | |
| | | *Day<sub>100</sub>* | -0.0051 | <0.001 | 0.0007 | $\pm 0.0014$ | | | |
| 2 | *k* | *AvAH* | -0.0022 | 0.0583 | 0.0011 | $\pm 0.0022$ | 0.3381 | <0.001 | 0.8966 |
| | | *Day<sub>100</sub>* | 0.0041 | <0.001 | 0.0006 | $\pm 0.0012$ | | | |
| 2<sup>†</sup> | *k* | *AvAH* | -0.0021 | 0.0643 | 0.0011 | $\pm 0.0022$ | 0.3259 | <0.001 | 0.7253 |
| | | *Day<sub>100</sub>* | 0.0047 | <0.001 | 0.0007 | $\pm 0.0014$ | | | |
| 3 | *k* | *AvTemp* | -0.0026 | 0.0020 | 0.0008 | $\pm 0.0016$ | 0.0853 | | 0.0045 |
| 4 | *k* | *AvAh* | -0.0042 | 0.0014 | 0.0013 | $\pm 0.0025$ | 0.0910 | | 0.0010 |
| 5 | *k* | *Day<sub>100</sub>* | -0.0045 | <0.001 | 0.0006 | $\pm 0.0012$ | 0.3280 | | 0.8427 |
| 5<sup>†</sup> | *k* | *Day<sub>100</sub>* | -0.0051 | <0.001 | 0.0007 | $\pm 0.0014$ | 0.3166 | | 0.3170 |

\* Adjusted $R^2$ for multivariable models
<sup>†</sup> Omits China (see text)

## Models: consequences and discussion

### U.S. states

We discuss the results of Model 4 in Table 1, which shows that variables *Day_100_* and log(*DP*) explain about 71% of the variability of the early daily growth rate estimate for U.S. states. The coefficient of *Day_100_* implies that if a state reached 100 cases 10 days after another one, one would expect *k* to be reduced by 0.08 point, which would impact significantly the pace of spread of Covid-19. For example, reducing *k* from 0.30 to 0.22, for two states with same population density value, the expected number of days to double the number of cases would increase from 2.3 to 3.2. Keeping those rates and starting from 100 cases, after one month the first state would have reached 810,000 cases, while the second, 74,000.

The value of the population density coefficient implies that doubling density would cause an expected increase of the value of *k* by 0.0212\**log*(2) = 0.0147 point. For example, the states of Iowa and Missouri, which reached the 100^th^ case on Mar. 23^rd^, had *k*’s estimated as 0.195 and 0.235, respectively, thus a difference of 0.040. Their population densities are 290 and 1,991 pop/k^2^, respectively, which imply an estimated increase of 0.041 point in the value of *k*, or almost exactly the actual difference. The scatter plots in Fig. 9 make it clear how *Day_100_* and *PD* variables are better associated to the variability of *k* than the weather ones.

The one-variable models for weather variables show an unexpected result, as both are positively associated to *k* in this case. But from Figs. 7.a-b, that behavior is in part caused by a late entry of the disease in colder and drier states, some with low population density values. But the results of Model 4 imply that the population density alone does not explain the variability of *k*. We further observe that, by themselves, the explaining power of *Day_100_* and population density, in relation to growth rate of Covid-19, are about the same (65% and 62%, respectively, Models 7,8). Thus, there are still other aspects impacting when state reached the 100^th^ case, along with population density (Model 9), possibly early measures of social distancing, many independent of government measures, such as people washing hands frequently, avoiding crowds and physical contact, restricting travels and circulation, etc., besides some societal and interpersonal contact practices. We see the timeline variable a likely candidate for a proxy of early social distancing initiatives and characteristics of societal and personal interactions in a region.

From Model 9, doubling the population density would make the day of 100^th^ case happen about 2.49*ln(2) = 1.7 days earlier. The scatter plot for these two variables (Fig. 10) shows that the states which had their outbreaks starting earlier, California, Massachusetts, New York and Washington, also show some outlier behavior in this case (especially the latter), as in the case of the association between *k* and the timeline variable (Fig. 9.c). The effect of population density on the date of 100^th^ case is expected, as not only lower population density slows the pace of contagion, as we have seen, but one may also think that the introduction of the virus would have been delayed in such states, due to lower traveling intensity to those areas (with the obvious exceptions of low-density areas with high level of tourist interest, as we have seen in the case of Idaho).

### Countries

According to Table 2, the date when the 100^th^ case occurred is the only significant variable for countries. Models 5/5^†^ say that, for each 10 days of delay in that occurrence, the value of *k* is expected to be reduced by 0.045 (0.051, for model without China), which is a smaller effect as in the case of U.S. states (0.080, Model 4, Table 1). This is likely due to the presence of outliers which we have not removed, like Japan and Singapore (see Fig. 11.c) and possibly others, which reduces the absolute value of the coefficient. Nevertheless, if we use the confidence interval for both coefficients, they cannot be considered statistically different.

Individually, weather variables show significative negative association with the pace of Covid-19 expansion, as already noted, a result that have been widely reported in other studies. In part, the reduced effect of the weather variables in the multivariable models including *Day_100_* could be explained by the plots and trend lines in Figs. 7.c-d, which show that the countries starting their outbreaks later showed higher temperatures and more humid weather. But, as we saw in the case of U.S. sates, the fact that weather variables may be associated with the date of the 100^th^ case cannot be used to reduce the impact of the timeline variable on the early growth rate of Covid-19, indicated in all multivariable regressions. Relaxing CI requirements, models 2/2^†^ show that, for countries with same date of 100^th^ case, increasing absolute humidity by 10g/m^3^ would reduce the value of *k* by about 0.02 point. That is a relatively small effect and would not severely impact the actual growth rate of Covid-19. Fig. 11 presents the scatter plots for *k* and control variables.

### Isolating the effect of population density

Using the results of Model 4 in Table 1, it is possible to remove the effect of the date when the state reached the 100^th^ case and estimate how population density impacts the initial daily growth rate of Covid-19 infection. From the value of the coefficient, one would expect that for every day we move a state from its original date of the 100^th^ case, *k* would increase by 0.008 point. We use the coefficient of *Day_100_* from Model 4 and bring all states back to March 7^th^, the date when Washington reached the 100^th^ case, the first U.S. state to reach that milestone. Following that methodology, we present, in Table 3, the results of the regression of the adjusted *k* w.r.t. population density. Figure 12 displays the scatter plot of the adjusted growth rate and population density, with the 95% CI band.

**Figure 12.**
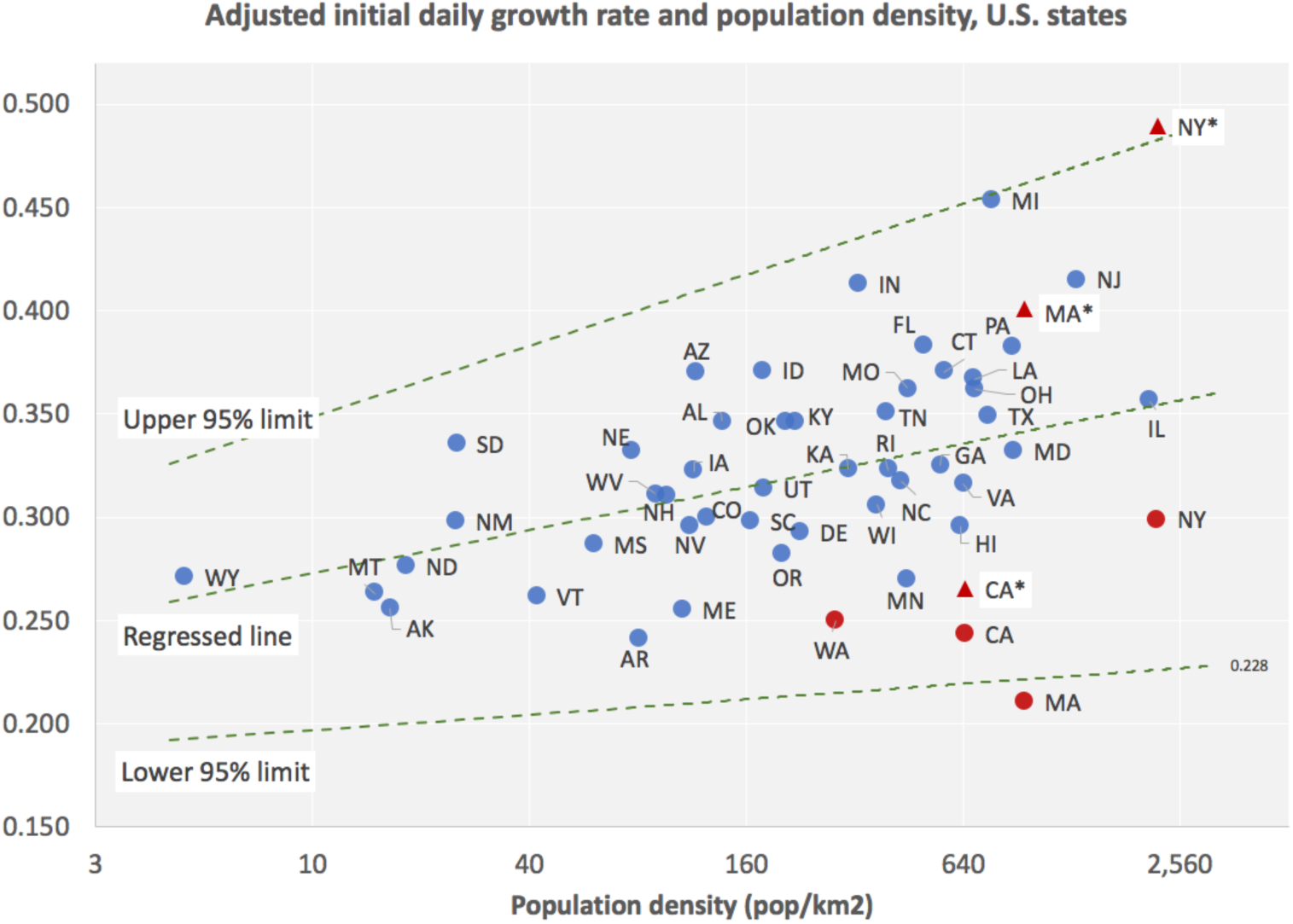
Adjusted initial daily growth rate and population density. 95% CI band between dashed lines. See text for the four states in red.

**Table 3.** Regression of adjusted *k* in terms of (log) population density, U.S. states.

| | Estimate | SE | p-value | Inferior<br>value (95%) | Superior<br>value (95%) | $R^2$ | Shapiro-Wilk<br>p-value |
| --- | --- | --- | --- | --- | --- | --- | --- |
| *Constant* | 0.2135 | 0.0224 | <0.001 | 0.1684 | 0.2585 | 0.3756 | 0.8350 |
| *Coefficient* | 0.0212 | 0.0041 | <0.001 | 0.0129 | 0.0296 |  |  |

The four states which were not part of the regressions employed along the process, including Massachusetts, are displayed in red. Disks use the adjusted values of growth rates, the triangles are adjusted for date of 100^th^ case from maximum values observed in the Figs. 3.a-c, the local maxima for California, Massachusetts and New York. In the case of Washington, as there is no local maximum in the graph for the evolution of k (Fig. 3.d), we did not perform any further correction, but it seems plausible, from the early behavior of the daily growth rate indicator, that the value we used for Washington is underestimated as well.

This is as close as we can get to estimate the relationship between a well-known independent variable and how Covid-19 would develop initially. We may now estimate the parameters *κ, α* in Eq. 1,

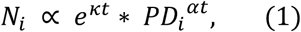

where *PD_i_* is the population density of U.S. state *i*. From the model in Table 3, one has

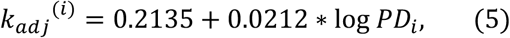

so that

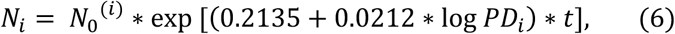

where *N*_0_^(*i*)^ is the number of cases on *Day_100_* for state *i*. Eq. 1 follows from (6) with *κ* = 0.2135, *α* = 0.0212. One would need to use the confidence intervals for these parameters, as given in Table 3, for a full estimate of the dependence of the initial evolution of cases on population density of state. In fact, one should include the intervals for the estimates of the adjusted *k* as well, which would introduce further uncertainty. The adjusted values and population density are plotted in Fig. 12, with 95% CI band. Any practical application of the above estimates regarding social distancing should be carefully confronted with actual field conditions and adjusted accordingly to actual response to adopted policies.

One would need to use the confidence intervals for these parameters, as given in Table 3, for a full estimate of the dependence of the initial evolution of cases on population density of state. In fact, one should include the intervals for the estimates of the adjusted *k* as well, which would introduce further uncertainty. The adjusted values and population density are plotted in Fig. 12, with 95% CI band. Any practical application of the above estimates regarding social distancing should be carefully confronted with actual field conditions and adjusted accordingly to actual response to adopted policies.

### Daily growth rate, population density and basic reproduction number

It is possible to relate the values of *k* to the *basic reproduction number* of an infectious disease, *R_0_*, the estimate of the average number of new infections generated by an infectious person (*31,32*), which has been subject of much investigation since the start of pandemic (*16*). With that in hand, Eq. 5 would provide the link to population density. For example, using a SEIR epidemiological model (*32*),

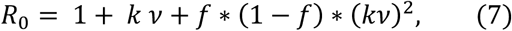

where *ν* is the serial interval and *f* is the ratio between latent period and serial interval, we see that, at least from an empirical viewpoint, *R_0_* depends quadratically on the logarithm of population density, since the daily growth rate shows good (empirical) linear dependence on density (Eq. 5).

For a state with low density, say 25pop/km^2^, the adjusted *k* would be in the interval [0.210, 0.354], and *R_0_*, in the interval [3.1,5.2], using the parameters in Eq. 7 from (*ν* = 7.5 days and *f* = 0.307) and our estimates for the daily growth rate without any social distancing in place. For a state with high density, say 1,600pop/k^2^, adjusted *k* would be in [0.264,0.477] and *R_0_*, in [3.8,7.3]. For New York City county, at 27,775pop/km^2^, if the model applies, one would have a range for *k* between 0.301 and 0.561 (the measured maximum was 0.511, see Fig. 3.c), and for *R_0_*, between 4.3 and 9.0. For Westchester, which has *PD* = 867pop/km2, the estimated interval for k would be [0.256,0.357] (the measured maximum was 0.345, Fig. 3.c), the interval for *R_0_* would be [3.7,7.0].

These values are higher than the ones estimated in (*33*) (baseline = 2.2), since they used a much lower estimate for the initial daily growth rate of Covid-19, of 0.10, estimated employing the very early phase of the disease in Wuhan, up to Jan. 4. Other estimates, using growth rates for a later period, much higher, have values closer to ours (*34*). These estimates have relevant implications for the amount of social distancing required to control the infection cycle. Basic epidemiological theory says that the force of infection should be reduced by 1–1/*R_0_* to achieve extinction. An *R_0_* of 3.1 would require a reduction of 71%; if it is 7.3, the required reduction would be 86%; if it is 9.0, 89%. These estimates are in line with what has happened in many places, as it is proving much harder to extinguish Covid-19’s infection cycle than originally estimated.

We observe that the above intervals for *R_0_* are possibly wider, as we would need to consider the intervals for *ν* and *f* as estimated in (*33*) and, as mentioned above, some extra variability coming from Model 4 in Table 1, used to compute the adjusted *k*. The above discussion is just a simple preliminary exercise indicating how population density enters the picture of estimating the basic reproduction number of Covid-19. Further studies regarding the estimation of parameters used to compute the basic reproduction number from initial daily growth rate would help establish its association with population density on firmer ground. One may also conjecture that there are underlining mathematical models directly relating *k* and *R_0_* to population density, which would provide better alternatives to establishing their relationship. Our empirical relationships could help check the adequacy of different models. Probably, we are just looking at first- and second-degree approximations of those relationships, which are reasonably adequate for the range of empirically available data.

The above models relating the early daily growth rate and basic reproduction number of Covid-19 to the variables of infection timeline and population density have shown similar results on a preliminary analysis of a set of U.S. counties with enough data and will be the subject of a follow-up paper (*35*), which will also discuss the case of Brazilian states.

## Final comments

Besides the results of the analysis developed in this paper, there is a group of U.S. states and countries with high levels of temperature and absolute humidity which have had high growth rate values in their early outbreaks (Table 4). Preliminary data for Brazilian states indicate that, like their U.S. counterparts, warmer and more humid weather have not precluded the expansion of Covid-19. For Amazonas (in the heart of the Amazon rainforest), Ceará and Pernambuco, states with fast early growth of Covid-19 cases, average temperatures were above 27C and absolute humidity levels above 20g/m^3^, from end of February to early April. Nevertheless, those states are undergoing faster outbreaks of the disease than Southern (colder/drier) states. Thus, any way one looks into the issue, it is hard to argue in favor of the idea that warm and more humid weather would help contain the spread of Covid-19.

**Table 4.** States/countries with *k* >0.200 and average temperature > 20C or average absolute humidity >10g/m^3^. *R^2^* of exponential regressions (Eq. 3), dates of 1^st^ (*Day_1_*) and of 100^th^ case (*Day_100_*).

| State/Country | Region | AvTemp | AvAH | $k$ | $R^2$ | Day1 | Day100 |
| --- | --- | --- | --- | --- | --- | --- | --- |
| Florida | USA | 23.9 | 13.4 | 0.319 | 0.972 | 2020-03-02 | 2020-03-15 |
| Brazil | Americas | 23.2 | 16.6 | 0.307 | 0.993 | 2020-02-26 | 2020-03-15 |
| Louisiana | USA | 21.7 | 14.1 | 0.295 | 0.982 | 2020-03-11 | 2020-03-16 |
| Ecuador | Americas | 19.8 | 10.1 | 0.291 | 0.974 | 2020-03-01 | 2020-03-18 |
| Texas | USA | 16.9 | 11.3 | 0.269 | 0.962 | 2020-03-05 | 2020-03-17 |
| Nigeria | SubSahAf | 29.1 | 20.2 | 0.261 | 0.981 | 2020-02-28 | 2020-03-31 |
| South Africa | SubSahAf | 18.3 | 10.1 | 0.259 | 0.993 | 2020-03-06 | 2020-03-19 |
| Thailand | Asia/Pacific | 29.4 | 19.1 | 0.248 | 0.942 | 2020-01-13 | 2020-03-16 |
| Alabama | USA | 17.7 | 11.0 | 0.243 | 0.985 | 2020-03-13 | 2020-03-20 |
| Malaysia | Asia/Pacific | 29.1 | 21.1 | 0.233 | 0.952 | 2020-01-25 | 2020-03-10 |
| Dominican Rep. | Americas | 23.7 | 17.9 | 0.220 | 0.887 | 2020-03-02 | 2020-03-22 |
| Chile | Americas | 21.1 | 8.8 | 0.220 | 0.992 | 2020-03-04 | 2020-03-17 |
| Saudi Arabia | MEast-NAfr | 21.5 | 5.4 | 0.220 | 0.977 | 2020-03-03 | 2020-03-16 |
| Panama | Americas | 29.9 | 18.5 | 0.219 | 0.993 | 2020-03-10 | 2020-03-18 |
| Indonesia | Asia/Pacific | 27.7 | 21.1 | 0.211 | 0.978 | 2020-03-02 | 2020-03-16 |
| New Zealand | Asia/Pacific | 17.8 | 11.2 | 0.210 | 0.979 | 2020-02-28 | 2020-03-23 |
| Argentina | Americas | 22.6 | 14.8 | 0.201 | 0.995 | 2020-03-04 | 2020-03-20 |
| Australia | Asia/Pacific | 21.7 | 13.6 | 0.200 | 0.994 | 2020-01-25 | 2020-03-10 |
| India | Asia/Pacific | 21.3 | 11.8 | 0.200 | 0.983 | 2020-01-30 | 2020-03-17 |

Northern hemisphere countries which will be enjoying Summer weather in the coming months, and warm and humid weather countries in Africa, Latin America, North Africa and Southeast Asia, should take that into account as they consider relaxing social distancing measures. It is possible that very high temperatures and levels of absolute humidity would help reduce the speed of Covid-19 spread, but the evidence from our study indicates that the best way to deal with Covid-19, at least until there are vaccines and/or effective therapeutics, taking into account the limitations of existing health systems, is to keep employing adequate levels of social distancing measures, whose results are predicted by modelling (*36*) and seem to be working effectively in all countries and states that have adopted them. The results on how population density relates to the growth rate of Covid-19 may help calibrate relaxation policies, with special care in the case of high-density communities.

Finally, our study suggests that the analysis of the dynamics of Covid-19 infection, or of any other similar infectious disease, should include population density considerations for subregions within a country or group of countries, which seem to allow for more detailed and complete understanding of the early dynamics of transmission and infection.

## Data Availability

Data files (csv) uploaded

## Acknowledgments

The author thanks Profs. Hildete and Aluísio Pinheiro of the Dept. of Statistics at Unicamp for discussions on the statistical aspects of this research. And Simon Schwartzman for observing that population density could be the actual relevant variable, sparking our interest in isolating its effect.

## Address for correspondence

Renato Pedrosa, Department of Science and Technology Policy, Institute of Geosciences, State University of Campinas (Unicamp). Rua Carlos Gomes, 250. 13085-855, Campinas, SP, Brazil;

